# Effect of COVID-19 pandemic on essential health care services and mitigation strategies implemented in Benishangul gumuz: An emerging region of Ethiopia

**DOI:** 10.1101/2022.11.16.22282389

**Authors:** Alemayehu Assefa Amaya

## Abstract

Health systems of low-to-middle-income countries may have fewer buffering resources and capacity against shocks from a pandemic. This study assessed the effects of COVID-19 on essential health care services and its mitigation strategies employed in Benishangul Gumuz Region in the Assosa district of western Ethiopia. Institutional based cross-sectional study design with multilevel triangulated mixed approach was used. Purposively selected health facilities and key informant or in-depth interviewee from different facilities were involved. Document review from health facilities reports and case registration logs were used to access the service utilizations per- and -post the pandemic. The study showed that there was a significant decrement in antenatal care (ANC) visits, family planning new acceptors, family planning repeated acceptors, skilled delivery, and in postnatal care within 7 days of delivery during the COVID-19 pandemic. There was an increase in home delivery, teenage pregnancy and still births. There was also a significant reduction in the vaccination coverage for almost all vaccines. During the pandemic, children treated for diarrheal disease and the number of people screened for human immunodeficiency virus (HIV), new HIV-positive individuals, and new HIV-positive individuals receiving antiretroviral therapy have decreased significantly. There has been a statistically significant increase in the number of people with elevated blood sugar during the pandemic. The COVID-19 pandemic has had a significant negative impact on essential health services, most notably in Benishangul Gumuz Region. Groups of individuals considered to be at higher risks of morbidity and mortality are the most significantly affected. Mitigation strategies have been implemented to address the indirect effects of the COVID-19 pandemic and improve health care delivery in the region’s highly affected areas. Urgent and comprehensive action is needed to mitigate the future (long-term) impact of Covid-19 on the health of community in the region.

## Introduction

Globally, efforts to control the spread of corona virus infection disease 2019 (COVID-19) have slowed the exponential growth of new cases, but have also led to unprecedented disorder in society, which has had a huge impact on health care, social, and economy (1,2,3). The pandemics has caused a concentrated nationwide response, during which other services, including the provision of healthcare have been neglected. The pandemics test the structure and competence of the health system. However, the post-pandemic period saw first-choice efforts to restore economic activity. The health system is still weak, at times getting weaker by the impact of the pandemic (4).

Currently, healthcare facilities in low- and middle- income countries (LMICs) are generally weak. It is stated that individuals in LMICs are deprived of adequate healthcare standard and cannot access medical facilities on time (5). In LMICs, even if there is no crisis, the coverage of essential health services is lacking, and they are often further overwhelmed and stretched during the pandemic. The challenges posed by the pandemic in LMICs are perhaps the most daunting. The pandemic has had an indirect impact on essential health services in many parts of the world, which may lead to increased morbidity and mortality and the loss of the gains made in the past few decades (6).

If this continues to be unmanaged, a strained health care system, essential health care interruption, and redistributed resources may lead to unrelated pandemic, maternal and neonatal morbidity and mortality, increased adolescent pregnancies and other reproductive health crisis are just like public health emergencies in the past. Evidence from the Ebola outbreak in Western Africa, for example, suggests that it has negative and indirect effects on sexual and reproductive health (SRH).

According to an analysis of data from Sierra Leone’s Health Management Information System (HMIS), decreases in maternal and newborn care due to disrupted services and fear of seeking treatment during the outbreak contributed to over 3,600 maternal deaths, neonatal deaths, and stillbirth (7). Evidence also shows that after the Ebola epidemic, the number of antenatal care visits and facility deliveries in Guinea did not recover to prior levels even after six months (8). This implies that the pandemics had sustained effects on the countries already inadequate level of health care.

Infectious diseases that require continuous support from the health system: Patients with tuberculosis (TB) and human immune deficiency virus (HIV) need a continuous supply of medicines. Interruptions in the intake of medicine are getting frequent. This is not only detrimental to the health of the patient but is also associated with the risk of the development of resistance to specific therapy. Correspondingly, a large number of patients of non-infectious chronic diseases (NCDs) including those who need periodic but regular administration of cancer therapies or long-term medication like diabetic and cardiac patients depend on medicines supply, monitoring and care of complications. Failure to provide these products and services may have serious negative impacts on the physical and mental health of these individuals (9).

The ratio of health professional to population in Ethiopia is 0.96 per 1000 populations (10), which is much lower than the world health organization (WHO) recommended standards, (4.45 per 1000 populations) to meet the sustainable development goal health targets (11). This suggests that the country’s health needs are not being adequately addressed and that an unplanned increase in COVID-19 related needs could exacerbate the country’s problems with essential health care services.

Based on these statements the COVID-19 pandemic could have critical effects on healthcare system, particularly in emerging regions like Benishangul Gumuz Region (BGR), where the health system is inadequate, poor infrastructure, extensive poverty, and high burden of other infectious disease like acquired immunodeficiency syndrome (AIDS), TB, malaria and neglected tropical diseases (NTDs) (12). However, there is no comprehensive scientific evidence on the impact of the COVID-19 pandemic on essential health services in Benishangul Gumuz Region. Therefore, this study aimed to assess the effects of COVID-19 pandemic on essential health services provision in Benishangul Gumuz Region in the Assosa district of western Ethiopia.

This will provide timely data and evidence to help in monitor and mitigate the crisis and assess the employed strategies to minimize its effects on essential health care services. It is also an opportunity to assess the existing health system and health infrastructure and response capacities during such crisis, which is important to assess continuity of essential services and provision of care during the crisis and in recovery. This will provide timely information on the status of essential health care services during the pandemic and post pandemic period which helps in discussion making.

In so doing this study, it will points policy makers to the way toward reforms that could improve the ability of the country and the region to cope with likely future epidemics and pandemics.

## Methods and Materials

### Description of the Study Area

For maps see fig. 1. Benishangul Gumuz Region is one of the ten Regional States of Ethiopia. The region is located in the western part of the country between 9° 30′-11° 30′ latitude in the north and 34° 20′-36° 30′ longitudes in the east with an average altitude of 1570m. The total land surface of the region is estimated to be 50,380 km^2^ (13). It is divided into three administrative zones: Assosa, Kamashi, and Metekel, and 20 districts or woredas. The regional capital city is Assosa. The region has three agro-climatic zones (13): the lowlands is the largest zone, located below 1500m altitude, representing more than 75% of the total area, with annual rainfalls in general lower than 400 mm. The second-largest zone is between 1500m and 2500m and represents 24% of the total area. The third and smallest zone (1%) is located above 2500m and includes mountain ranges and high plateaus.

The regional capital, Assosa is located at a distance of 687 km away from the capital city of the country, Addis Ababa. The region has also an international boundary with Sudan and South Sudan on the west, and it is bordered by the Amhara Regional State on the east and north, by the Oromia Regional State on the east and southeast, and by the Gambella Region on the south (13).

Assosa district is one of the 3 districts in Benishangul-Gumuz Region of Ethiopia. The majority ethnic group in the zone is the Berta people, and it is also home for different ethnic groups from different Ethiopian regions. The zone has two hospitals (one general and the other primary) and 23 health centers of which only 9 of them providing all essential health care services.

### Study Design and Approach

An institutional based cross-sectional study design with multilevel triangulation-mixed method approach involving quantitative and qualitative data was implemented to assess the effects of COVID-19 pandemic on essential health care services before and after the COVID-19 pandemic and to evaluate response measures taken against the effect of the pandemic on essential health care services in Benishangul Gumuz Region in the Assosa district of western Ethiopia. It is multilevel, because it involved different stockholders (individuals, health care providers at different essential health care unit and program implementers at different level). It is triangulation mixed, because, different, but complementary data to answer similar research objectives were used.

### Source and Type of Data

Secondary data (monthly reports on essential health care services) from selected health facilities (Assosa general hospital, Assosa HC, Bambasi health center, Homosha health center, Kurmuk health center, Menge health center, Menge Hospital, Sherekol health center and Buldigilu health center) and primary data through interviewing the following concerned bodies from: Regional health bureau (RHB) representative, maternal and child health officer, NCD officer, HIV officer, NTDs officer, malaria officer, immunization officers, logistic officer and Pharmacal head, and the following concerned bodies from each health facilities: facility head, finance head, TB focal person, TB patients, HIV focal person, people living with human immunodeficiency virus (PLHIV), immunization focal person, families vaccinating their child, outpatient diagnosis (OPD) focal person, antenatal care (ANC) focal person, deliver unity focal person, laboratory head, family planning (FP) focal person, malnutrition management center focal person, and Pharmacy head using chick lists. observations were also made at selected health service centers.

### Sample Size Determination and Sampling Procedure

The sample size of the study was nine health facilities, the health facilities actively giving essential health care services in the zone and 24 interviews were planned. Purposive sampling technique was implemented to select health facilities and the individuals for interview. If two or more health personnel are found at the service centers, one health personnel were selected by simple random sampling. Similarly, if two or more health service seekers or patients are found at the service centers, one was selected by simple random sampling

### Eligibility criteria

All health care facilities in Assosa district that give essential health care services during the last four years and that report using HIMS under the jurisdiction of the regional health bureau were included in the study.

### Data Collection Tools and Techniques

Data was abstract from monthly essential health care reports from each health facilities using standardized forms for data gathering and the indicated target persons were interviewed using chick lists. For data abstraction from reports, reports were categorized in to two sections; before the pandemic (July, 01,2018 to December,30,2019) and after the pandemic (January,01,2020 to June 2021) in Ethiopian. Data extraction standardized form, structured/semi-structured questioners and interview guides were used. The Participants were asked a series of questions regarding the effects of COVID-19 pandemic on healthcare system and the response measures taken against the effect (workers safety support related responses, patient service delivery related responses, and data streams for situational analysis).

### Data Management and Analysis

#### Quantitative Data Management and Analysis

Quantitative data were analyzed using SPSS version 25. Descriptive statistics are used to summarize distribution frequencies and are presented using a table. The Wilcoxon signed-rank test was used to compare observations between the peri-COVID-19 period and the post-COVID-19 pandemic period, with a significance level of α = 0.05.

#### Qualitative Data Management and Analysis

The qualitative data was analyzed manually using the thematic method as previously described (Braun *et al*., 2006). All qualitative data was collected by audio-recording, and the verbatim responses to each question were translated and transcribed, and recorded in Microsoft Word. A code sheet has been created following the focus group and the key informant guides after which the textual data were encoded into selected themes and a master sheet analysis was carried out. Thematic analysis is used to categorized into themes and then formulate ideas by looking at the response patterns. The analyzed data was presented in text form.

### Data Quality Assurance

The team was oriented and trained for two days on basic techniques. They were supervised by experienced health professionals and the filled questionnaires were checked daily. Furthermore, the questionnaire was pre-tested. In addition, to assure the quality of qualitative data, two individuals were independently immersed themselves in the data within the highlighting process. The two individuals then meet to determine codes that accurately represented the participants’ experiences and define each theme. Then, two people independently encoded (line-by-line) the complete responses from all participants to establish an inter-rater agreement. Coding disagreements were resolved through discussion and review of the data.

## Results

Nine health facilities in Benishangul Gumuz Region in the Assosa district (including 7 health centers and two hospitals) were included in this study. From planned 24 interviews only 14 Interviews were conducted due to information saturation on this number.

### Effects of COVID-19 pandemic on maternal, neonatal and child health care services

The results of this study show that the hard lockdown during the COVID-19 pandemic has greatly affected access to and utilization of essential healthcare services in the Benishangul Gumuz Region. The study showed that there was a significant decrease of 29%,50%,15%,32.2%, and 80.5%, in at least 4 ANC visits, family planning new acceptors, family planning repeated acceptors, skilled delivery, and in postnatal care within 7 days of delivery during the COVID-19 pandemic compared to the pre-pandemic period respectively (Table 1).

There was an increase in the number of home birth notices (42%), teenage pregnancies (38.2%) and stillbirths (36.4%). The increase in the number of home birth notices was statistically significant (P=0.008), but the increment in the number of teenage pregnancies and stillbirths was not statistically significant (Table 1).

No significant differences were observed in neonatal deaths within 7 days of delivery (institutional), neonates treated for sepsis, and notified neonatal deaths (community based) before and after the COVID-19 pandemic (Table 1).

There was a decrease in BCG vaccine doses given (27.5%) which is significant (P=0.033) and a decrease in polio vaccine doses given (49.4%), pentavalent vaccine doses given (47%), and pneumococcal conjugated vaccine doses given (47%) which are all significant (P=0.043). The number of measles vaccine doses given (all ages) was also come down by (53.8%) which is significant (P=0.014) (Table 1).

There was a significant (P=0.001) decrease in the number of children treated for diarrhea (124%) during the COVID-19 pandemic compared to the per pandemic period (Table 1). A clinician at under 5 OPD in one health facility said that “…*the number of children presented with complaint of diarrhea was highly reduced during the pandemic, but later after the pandemic peak the diarrhea cases are increasing…*”

The study found that there were no significant excess overall still births and community based neonatal death notification in the post-pandemic compared to the pre-pandemic period, although the number of causes were higher than the level of per-pandemic period (Table 1). A pregnant mother coming for ANC at one health facility said that “the *time was stressful; it was not easy to visit health facilities due to fear of the pandemic and the health professional were also in stress during the time… many interrupt their ANC follow up and home delivery was also common…”*

The study also found that there was no significant decrement in number of under 5 children screened for malnutrition and diagnosed as malnourished post-pandemic period. Health professional at under 5 OPD in one health facility said that “*generally the number of malnourished children is increasing, post-pandemic the number of children coming to our facility is decreased, but the number of malnourished children is increasing*…”

### Effects of COVID-19 pandemic on HIV and sexually transmitted infections care services

There was a decrease of 42%,129% and 106.5% in the number of individuals screened for HIV, new HIV positive individuals, and new HIV positive individuals put on ART during the pandemic compared to pre-pandemic period respectively. The decrements are statistically significant at p-value of 0.043, 0.012 and 0.035 respectively (Table 2).

The COVID-19 pandemic has imposed a great economic and psychosocial impact on people living with the virus. A woman on ART treatments coming for collecting her ART medication at one health facility said that “the *media, the community, and even the health professionals are talking that we are the high-risk group and this makes us hopeless and highly stressed. We are stigmatized, we lost our freedom, and it was difficult to access our medications. Personally, I have missed many pills, but later the health professional extends the medication collection time to 6 months which actually reduced our suffering*.*”*

Social isolation and Loneliness were also another challenge for PLHIV. A woman at ART clinic said that *“I was separated from my families and my friends and this condition worsen my stress. There is a time that I diced to hung myself…anyways it is passed thanks to GOOD. However, we are still in financial crisis …”*

Morbidity and mortality among PLHIV were high and the number of HIV testing service seekers were highly reduced during the pandemic. For example, a health professional working in ART clinic at one health facility said *“the period was stressful especially to PLHIV and health professionals working in ART clinic… most patients have missed their appointment and their pills. Most of PLHIV were severely affected by the pandemic and most are mentally disordered during the pandemic. HIV test and prevention measures were highly affected during the pandemic, because the numbers of service seekers were highly reduced due to fair of COVD-19; health professionals fair of the pandemic; shortage of test kit and personal protection equipment and materials*.

Adherences to ART has been greatly reduced during the pandemic and the ART drop rate has been high, and HIV incidence has also been greatly reduced during the pandemic. For example, a health official said that *“the effect of the pandemic on adherences and ART follows up, Viral load test and other follow-ups and health services was high. For example, about 170 PLHIV was lost from follow-up during the pandemic, who were later traced and about 80% of them returned to the regular follows up*. …*the numbers of new HIV cases has been greatly reduced during the pandemic, this is actually due to the numbers of people tested for HIV during the pandemic were low comparatively”*

There was a decrease of 93%,82.4% and 88% in the number of individuals screened for sexually transmitted infection (STI) by syndrome diagnosis, STI cases treated, and STI cases partners traced and treated during the pandemic compared to pre-pandemic period respectively. The decrease is statistically significant at a p-value of 0.028 (Table 2).

An interview with a clinician working at OPD also confirms the decrement in the number of people visiting health facilities for STI screening and the STI cases during the pandemic. He said that “*health facilities visiting for non-COVID-19 related cases was actually reduced during the pandemic and similarly individuals seeking STI care services was reduced and the incidences of STI cases was also reduced. I don’t think social distancing measures really reduced STI incidences, rather it is probable due to cases are not seeking services*.*”*

### Effects of COVID-19 pandemic on TB and malaria care services

There was no statistically significant increase in the number of people screened for TB (5.5%), but there was a statistically significant decrease in the number new TB cases (131.6%) during the pandemic compared to the pandemic period (P=0.042) (Table 3).

However, COVID-19 pandemic has imposed a great economic and psychosocial impact on old tuberculosis cases. A man on TB treatment coming for collecting his anti-TB medication at one health facility said that “*the media, the community, and every one talking that TB patients are at higher risk for COVID-19 infection and at high-risk of death. This leaves us in despair. We are stigmatized, we lost our freedom, and it was difficult to access our medicine. Personally, I missed many pills, but latter the health professional gave us our medication for 4/6 months which reduced our suffering*.*”* A health professional working in a TB clinic at one health facility said *“the period was stressful especially to TB patients and health professionals working in TB clinic… most patients have missed their appointment and their pills. Later after the peak pandemic time was passed, we tried our best to trace the missed patients and make their refilling time longer and also have provided personal protections and psychosocial supports. The number of new cases is reduced…”*

The number of people visited health facilities for malaria and the number of individuals treated for malaria was decreased by 13.3% and 22.7% respectively, which is statistically significant at p-value of 0.043 and 0.022 respectively (Table 3).

### Effects of COVID-19 pandemic on non-communicable disease care services

There was no a statistically significant increase in the number of individuals screened for DM (41%), but there was a statistically significant increase in the number of people with raised blood sugar (74.4%; P=0.041) during the pandemic compared to pre-pandemic period (Table 4).

The number of people visited health facilities for HTN screening (56.8%) and the number of individuals with raised blood pressure (73.4%) was significantly increased at p-value of 0.032and 0.006 respectively (Table 4). The high number of screened HTN risky people makes the number of people with raised blood pressure higher in this study.

A hypertensive woman coming for follow-up at one health facility said that “I don’t want to remember that time, *I was waiting day of my death and this makes us hopeless…I was stress much, isolated from my families… the other challenge was increase in price of medication and inaccessibility which worsen our problem…*

### Impacts of COVID-19 on Neglected Tropical Diseases (NTDs)

In similar way as other health system COVID-19 have had great impact on NTDs control and elimination efforts. An official working in the program said that “*… have highly affected the activities of NTDs case management, deworming campaign and other prevention measures activity… for example, deworming campaigns were missed due to budget shortage or shift, human power shortage/shift, movement restriction due to the pandemic. Health facilities visiting was reduced and one of the NTDs controlling strategy treating cases was missed. These all affects the NTDs prevention and control efforts*.*”*

### Financial impact to health care system due to COVID-19

Many health facilities reported experiencing financial instability because of increased expenses associated with responding to the pandemic and lower revenues from decreased use of other health facilities services. For example, a finance officer at one health facility said that “*there was shortage of supplies due to budget shift to COVID-19 prevention and control activities, and financing from NGOs was also reduced and transportation and movement restriction. Patient flow for other health care services also have been reduced and because of that the revenues from services is simultaneously reduced. We have been facing high shortage of medication especially for chronic disease like diabetes, heart disease and hypertension*.*”*

Unlike to urban health facilities that reported both patient fairs to come to health facilities and shortage of supplies as primary for redaction in essential health care services, rural health facilities reported shortage of supplies as the primary reason.

### The Health Systems Impact of COVID-19 mitigation strategies undertaken

Transport and border restrictions introduced by community quarantine measures have been universally impacted health services access and delivery globally. This study showed that accesses to most basic health care services and health facilities come down during the pandemic (Table 1-4). Qualitative data collected through interview indicated, care seeking behaviors in both providers and patients have also changed as a result of restrictions and the fear of contracting COVID-19. On the supply side, the measures to contain COVID-19 have been drain off away significant manpower and resources that provide routine essential services.

Mitigating measures for sexual and reproductive health services disrupted due to the pandemic are; giving services by adhering to COVID-19 prevention precautions, phone-based counseling and support, extending appointment durations for normal pregnancies, and community outreach mobilization and tracing after the peak of the pandemic are among the most common mitigation measures undertaken.

For example, a health professional working in MCH center in on health facility said that “…*during the pandemic most of the time the cases visiting our health facility were high risk individuals, like individuals who are twin pregnancy or other obstetric cases…the rate of stable cases visiting the facility have been significantly reduced. During the peak pandemic period we stayed providing services for those coming to the facility under restrict COVID-19 prevention protocol, but later after the peak the pandemic we implemented tracing and community outreach strategies and know it is much better*.*”*

Mitigating measures undertaken to reduce the possible impact of COVID-19 pandemic on HIV/AIDS care and services were the flowing as obtained from different concerned bodies at different health facilities. Home based medication supply through using health extension worker, extending the duration ART refiling and appointment 6 months for stable HIV cases and for every 2-3 months for new unstable HIV cases, overlapping all HIV/AIDS related services for PLHIV, shorting stay hours in the health facilities and in addition providing self-protection materials (facemask, glove, sanitizer) and early awareness creation on COVID-19 vaccine and giving priority to vaccinate this group of people are the major ones. A health official working on the program said that *“…. we tried our best through these mitigation methods to minimize the impact of the pandemic on HIV/AIDS services and the prevention and control of the transmission. By doing this we able to traced about 80% of lost PLHIV and we hope the probability of COVID-19 infection among this group of people is reduced due to our mitigation measures*.*”*

Mental health problems (stress, being panic, depression) are common among the community, especially among individuals thought to be at high risk of morbidity and mortality of Covid-19 per information collected from health professionals and services seekers at different center of the health facilities. Dispute the reality of increment in mental health care service there was no additional new mental health services centers or improved capacities in the region. For example, a health person in one health facility said that “*…health seeking behavior for mental health is poor in the region and there is no strong mental health services center in the region and no new related activities done related with COVID-19 pandemic…actually mental health problem is raised especially among individuals thought to be high risk for the pandemic”*

A woman collecting here ART at one health facility said that “…most of our friends were mentally affected, we were lonely for long time and when we are alone, we think all the time about the condition. It was also difficult to go out for work and we lost our income. This all makes us stressed and depressed. Personally, I was sleepless for several months and there was a time I missed meaning of life… we need supports.”

This study has strength and limitations. The study includes both quantitative and qualitative data, so the weakness of one type of data is optimized by triangulation of the other type of data. Despite this strength, since the main source of data for this study is secondary data from health facilities report and HIMS whose data are at times not good before validation exercise and the limited number of grouping variables. Such accumulated data are known to have high levels of inconsistency, missing values, and invalid records and this may be worse during the COVID-19 pandemic and in low-resource settings like Benishangul Gumuz. Secondly, the analysis cannot fully attribute the changes to intervention because of lack of control or counterfactual data. The intervention affected the whole country and thus there were no areas that could work as control; thus, stronger analytical techniques like controlled interrupted time series (CITS) analysis cannot be applied. These limitations were overcome through taking only completed data and through data triangulation.

## Discussions

The study showed that there was a significant decrease in ANC visits, family planning new acceptors, family planning repeated acceptors, skilled delivery, and in postnatal care within 7 days of delivery, and a significant reduction in the vaccination coverage and decline in total number of vaccines administered during the COVID-19 pandemic compared to the pre-pandemic period respectively. There was an increase in the number of home delivery notices, teenage pregnancies, and stillbirths. There was also a significant decrease in the number of children treated for diarrhea, people screened for HIV, new HIV positive people and the new HIV positive individuals put on ART, people screened for STI by syndromic diagnosis, STI cases treated and the STI cases partners traced and treated during the pandemic compared to pre-pandemic period. Many health facilities also reported experiencing financial instability because of increased expenses associated with responding to the pandemic and lower revenues from decreased use of other health facilities services.

The study showed that there was a significant decrease in ANC visits, family planning new acceptors, family planning repeated acceptors, skilled deliveries, and postnatal care within 7 days of delivery during the COVID-19 pandemic compared to the pre-pandemic period respectively. This is similar to a study in Ethiopia (14) and it is higher than 10% redaction predicated for low-income countries (15).

This is might be due to a strained health care system, disruption in care, and redistribution of resources and fear of service seekers to visit health facilities (7). As it is stated below, this leads to high home delivery rate, high in still birth rate and teenage and unwanted and unplanned pregnancy. For example, a woman at ANC center come for ANC service said that “I *have six children and I planned to be more pregnant and I was using injectable form of contraceptive, but I have missed my appointment day during the pandemic. Because of it I become pregnant (unwanted and unplanned) …”*

No significant differences were observed in neonatal deaths within 7 days of delivery (institutional), neonatal deaths reported (community-based), and neonates treated for sepsis during the pre-pandemic and post-pandemic period. This is likely due to the fact that the community has not reported all cases, and as noted above, the number of institutional deliveries has decreased significantly during the pandemic.

There was a significant decrease in almost all vaccine doses given. This finding supports the United Nations report of COVID-19 negative effect on vaccination (16). A reduction in the vaccination coverage and decline in total number of vaccines administered, led to children missing out on their vaccine doses is also reported by a systematic review study (17). This may put the region at risk of vaccine preventable diseases outbreaks, extremely contagious diseases like measles (18)

There was a significant decrease in number of children treated for diarrhea during the COVID-19 pandemic compared to the pre-pandemic period. A clinician at under 5 OPD in one health facility said that “…*the number of children presented with complaint of diarrhea was highly reduced during the pandemic, but later after the pandemic peak the diarrhea cases are increasing…*” No similar research works were identified during literature search. This low diarrheal disease incidence rate may be due to adherence to the COVID-19 preventive measures and protocols could be protective and reduce the incidence diarrheal disease (19).

The study found that there were no significant excess overall still births and community based neonatal death notification in the post-pandemic compared to the pre-pandemic period, although the number of causes was higher than the level of pre-pandemic period. Higher number of still birth and community based neonatal death is what is expected because of significantly reduced ANC follow-up and post-natal care follow-up, both causing home delivery (15). The non-significant increment cases may be due to non-reporting all cases due to restriction of movements, fear of the pandemic and/or the parents not need to report the cases and the health professional not tracing the cases. A pregnant mother coming for ANC at one health facility said that “the *time was stressful; it was not easy to visit health facilities due to fear of the pandemic and the health professional were also in stress during the time… many interrupt their ANC follow up and home delivery was also common…”*

The study also found that there was no significant decrease in the number of under 5 children screened for malnutrition and diagnosed as malnourished during the post-pandemic period. This is due to decrement in number of children visiting the health facilities during the pandemic. However, the prevalence of malnourished individual among children was higher post-pandemic compared with pre-pandemic period. Health professional at under 5 OPD in one health facility said that “*generally the number of malnourished children is increasing, post-pandemic the number of children coming to our facility is decreased, but the number of malnourished children is increasing*…” This finding is agreed with a study report by a Standing Together for Nutrition consortium (ST4N) (20). This may be due to steep declines in household incomes, changes in the availability and affordability of nutritious foods, and interruptions to health, nutrition, and social protection services, especially among poor households leading in difficulty in accessing food (21,22, 23)

The COVID-19 pandemic has imposed a great economic and psychosocial impact on people living with the virus. A woman on ART treatments coming for collecting her ART medication at one health facility said that “the *media, the community, and even the health professionals are saying that we are the high-risk group and this makes us hopeless and highly stressed. We are stigmatized, we lost our freedom and it was also difficult to collect our medicines. Personally, I have missed many pills, but later the health professionals had extended our medication collection time to 6 months which reduced our suffering*.*”*

Social isolation and Loneliness were also another challenge for PLHIV. A woman at ART clinic said that *“I was separated from my families and may friends and this condition worsen my stress. There is a time that I diced to hung myself…anyways it is passed thanks to GOOD. However, we are still in economic crisis …”* This is a similar finding with study reported by similar study elsewhere *(24)*.

Morbidity and mortality among PLHIV were high and the number of HIV testing service seekers have been highly reduced during the pandemic. For example, a health professional working in ART clinic at one health facility said *“the period was stressful especially to PLHIV and health professionals working in ART clinic… most patients have missed their appointment and their pills. Most of PLHIV were the most affected and most are mentally disordered during the pandemic. HIV test and prevention measures were highly affected during the pandemic, because the numbers of service seekers were highly reduced due to fair of COVD-19; health professionals fair of the pandemic; shortage of test kit and personal protection equipment and materials*.

Adherence to ART is greatly reduced, and ART drop rate are high. During the pandemic, the incidence of HIV has been greatly reduced. For example, a health official said that *“the effect of the pandemic on adherences and ART follows up, Viral load test and other follow-ups and health services was high. For example, about 170 PLHIV was lost from follow-up during the pandemic, who were later traced and about 80% of them returned to the regular follow-up*. …*the number of new HIV cases has dropped significantly during the pandemic as very few people are tested for HIV*.*”*

The interruption of HIV/AIDS health care services during the pandemic is also reported by similar studies indifferent parties of the world (22,23). World health organization African region indicated that reduced quality clinical care owing to health facilities becoming overstretched, and a suspension of viral load testing, reduced adherence counseling and drug regimen switches (25). This disruption in treatment and prevention measures may results in increased morbidity, mortality, HIV transmission and drug resistance, which highly affects the prevention and control efforts of HIV/AIDS.

There was a significant decrease in the number of people screened for STI by syndromic diagnosis, STI cases treated and STI cases partners traced and treated during the pandemic compared to the pre-pandemic period respectively. This redaction in STI case reporting was also reported in a similar study elsewhere (26). This fact can be explained with significant underdiagnosis and underreporting of cases during the pandemic because of reduced screening or it also may reflect a true decline in STI transmission due to social distancing measures during the pandemic (27). This disruption in treatment and prevention measures of STIs may results in increased morbidity, STI transmission, HIV transmission and drug resistance, which highly affects the prevention and control efforts of STIs and HIV/AIDS (27).

There was no statistically significant increase in the number of people screened for TB, but there was a statistically significant decrease in the number of new TB cases during the pandemic compared to pre-pandemic period. These findings are similar to a study in Japan and India that reported a significant decrease in new-causes TB after the COVID-19 pandemic (9). This is may be due to reduction in the prevalence of other infectious diseases due to greater using of Personal Protective Equipment (PPE), social distancing and other sanitizing practices during the pandemic (19,28).

However, COVID-19 pandemic has imposed a great economic and psychosocial impact on old tuberculosis cases. A man on anti-TB medication at one health facility said that “*the media, the community, and every one talking that TB patients are risky group for COVID-19 infection and this makes us stressed. We are stigmatized and we lost our freedom. Personally, I missed many pills, but later the time duration of medication collection was extended to 4/6 months, which reduced our suffering*.*”* A health professional working in a TB clinic at one health facility said *“the period was stressful especially to TB patients and health professionals working in TB clinic… most patients have missed their appointment and their pills. Latter after the peak pandemic time was passed, we tried our best to trace the missed patients and make their refilling time longer and also have provided personal protections and psychosocial supports. The number of new cases is reduced…”*

However, the new TB cases reduced due to COVID-19 protection protocols, the disruption in treatment and treatment drop rate may results in increased morbidity, mortality, TB transmission and drug resistance in M. TB, which highly affects the TB prevention and control efforts.

The number of people visited health facilities for malaria investigation and people treated for malaria was significantly reduced. This finding is similar with studies from malaria endemic countries that reported the redaction of people screened for malaria and malaria cases treated in the health facilities (29). This redaction is likely due to reductions in malaria screening and treatment in health facilities due to the fear of contact with COVID-19 cases and the lack of transportation access due to community quarantine. The redaction in malaria screening, treatment and other prevention measures might have increased morbidity, mortality and malaria transmission that have great negative impact on malaria prevention, control and elimination efforts of the region as well as national (30).

There was no statistically significant increase in the number of people screened for DM, but there was a statistically significant increase in the number of people with elevated blood sugar during the pandemic compared to the pre-pandemic period. Bill published a similar report explaining the possible cause of elevated blood sugar levels (DM) during the COVID-19 pandemic (31). Nonetheless, this requires further investigation, but in this study, individuals at higher risk for DM were screened, possibly due to fear of higher risk for COVID-19.

The number of people visited health facilities for HTN screening and the number of individuals with raised blood pressure was significantly increased. In this study, the increment is due to individuals at higher risk for HTN were screened, possibly due to fear of higher risk for COVID-19. It may also be due to increased blood pressure due to COVID-19 panic among people at risk for of high blood pressure (32).

The impact of COVID-19 on the follow-up and care of chronic diseases like diabetes and hypertension is similarly reported in a narrative review describing as “the lockdown of different services decreased referral, access and hospitalization resulting in inadequate ongoing care for chronic conditions among needy patients” (33).

This due to the reallocation of resources from different sources from chronic disease prevention, diagnosis, management, and rehabilitation during the outbreak. The lockdown of different services has also reduced referrals, access and hospitalizations, leading to inadequate ongoing care for chronic conditions for patients in need (34).

Many health facilities have reported financial instability due to increased costs associated with responding to the pandemic as well as reduced revenue due to reduced use of other health facilities services. Unlike urban health facilities reporting patients fear to come to health facilities and supply shortage as the main reason, rural health facilities reported supply shortage as the main reasons.

## Conclusions and Recommendations

The study shows that the COVID-19 pandemic and its responses, such as lockdown and community quarantines, have had a significant negative effect on essential health services, most notably in Benishangul Gumuz Region in the Assosa district of western Ethiopia. Groups of individuals to be at higher risk of morbidity and mortality are the most significantly affected. Some sort of strategy for the indirect effects of the COVID-19 pandemic, as health officials explained in interviews, has improved the district highly affected health care delivery.

The researcher recommends an urgent and comprehensive action to mitigate the future (long term) impacts of Covid-19 on health of the community such as: treatment adherences and drug resistance, worsening of chronic conditions, disease outbreaks, maternal and child mortality, chronic undernutrition, and the possibility of an increment in infection and mortality from HIV/AIDS/STI. This raises the need for global collaboration in national planning, health system strengthens, community-based measures, and donor investments to prevent the reintroduction and re-establishment of existing endemics to avoid increased morbidity and mortality. Protecting the long-term health outcomes and improving the financial security of groups of people living in deprivation is also strongly recommended for local and national concerned government bodies and non-governmental organizations working in the area.

## Data Availability

Data used in this manuscript will be provided up on request

## Declarations

### Abbreviations

AIDS: Acquired immune deficiency syndrome
ANC: Antenatal Care
ART: Antiretroviral therapy
BGR: Benishangul Gumuz Region
CDC: Communicable diseases control
COVID-19: Coronavirus disease 2019
DM: Diabetes Mellitus
EIDs: Emerging infectious diseases
EVD: Ebola virus disease
FP: Family Planning
HCPs: Health care providers
HCWs: Health care workers
HF: Health Facilities
HIV: Human immunodeficiency virus
HTN: Hypertension
KI: key informant
NCD: Non-communicable Disease
OPD: Outpatient Diagnosis
PLHIV: People living with human immune deficiency virus
RHB: Regional health Bauru
STIs: Sexual Transmitted Infections
UNAIDS: United Nations AIDS found
WHO: World Health Organization

## Acknowledgments

I would like to thank the study participants for voluntarily taking part in the study for their participation and local health officials and community leaders for their cooperation. I also thank data collectors for their commitment and collecting quality data.

## Author contributions

All core activities: conceptualization, filed work and data collection, formal analysis and investigation, and result writing and manuscript preparation were done by the author [Alemayehu Assefa].

## Availability of data and materials

All data that support the findings of this study are available from the author upon request

## Ethics approval and consent to participate

The planning, the conduct, and reporting of the research are in accordance with the Helsinki Declaration. The study was approved by the Institutional Review Board of Assosa University (Ref No: ASU/IRB/003/2017/22). Prior to conducting of the study, a letter of support was obtained from the district health offices and meetings were held with community leaders and community members to explain the aims of the study. The aim of the study was explained to the study participants, and an informed written and verbal consent have been obtained from each participant Ethical approval and informed consent forms and paper copies of signed consents have been kept in a confidential place.

## Consent for publication

Not applicable

## Supporting information files

S1 Fig: Map of the study area. Source: BGR,2020

S1 Table: Effects of COVID-19 pandemic on maternal, neonatal and child health care services in BG, west Ethiopia

S2 Table: Effects of COVID-19 pandemic on HIV and STI care services in BGR, Assosa Zone, west Ethiopia

S3 Table: Effects of COVID-19 pandemic on TB and malaria care services in BGR Assosa zone, west Ethiopia

S4 Table: Effects of COVID-19 pandemic on non-communicable diseases care services in BGR Assosa zone, west Ethiopia

